# Genetic liability to bipolar disorder and body mass index: A bidirectional two-sample mendelian randomization study

**DOI:** 10.1101/2022.01.31.22270154

**Authors:** Lars Meinertz Byg, Maria Speed, Doug Speed, Søren Dinesen Østergaard

## Abstract

**Objectives:** Bipolar disorder is associated with increased body mass index (BMI), but it remains undetermined if this association is causal and, if so, in which direction it goes. Here, we sought to answer these questions using bidirectional two-sample Mendelian Randomization, a method from genetic epidemiology that uses data from genome-wide association studies (GWAS) to examine whether a risk factor is causal for an outcome.

**Methods:** We used summary statistics from GWAS of bipolar disorder and BMI conducted using data collected by the Psychiatric Genomics Consortium and the UK Biobank, respectively. The genetic instrument for bipolar disorder contained 53 SNPs and explained 0.5% of phenotypic variance, while the genetic instrument for BMI contained 517 SNPs and explained 7.1% of phenotypic variance.

**Results:** We found that a two-fold increase in the genetic liability to bipolar disorder causes a 0.6 (kg/m^2^) reduction in BMI, which is statistically significant (P=0.009), and predominantly driven by reduced fat mass. Conversely, we found no evidence that BMI causes changes in the risk of developing bipolar disorder (P=0.59).

**Conclusion:** The findings of this study indicate that the increased BMI observed among individuals with bipolar disorder is not a direct consequence of genetic liability to bipolar disorder, but more likely represents the sum of downstream correlates of manifest bipolar disorder, such as side effects of pharmacological treatment, poor diet and sedentary lifestyle, that are all modifiable and could potentially be targeted as part of clinical management.

## Introduction

Bipolar disorder is associated with a substantial reduction in life expectancy [1, 2]. Of the many contributors to this association, obesity is likely one of the most important [3, 4]. This is because obesity is known to increase mortality [3-5] and there are many studies linking bipolar disorder to obesity [6]. However, it remains unknown whether this link is a direct causal effect of the genetic liability to bipolar disorder. Using Mendelian randomization, we recently demonstrated that for depression, a disorder which shares both phenotypical traits [7-9] and genetic risk [10] with bipolar disorder, obesity appears to be a causal risk factor (higher BMI causes higher risk for depression) and not vice versa [11], which speaks against bipolar disorder being causally related to increased body mass index (BMI).

Mendelian randomization is a method from genetic epidemiology that uses data from genome-wide association studies (GWAS) to examine whether a risk factor is causal for an outcome [11-15]. Specifically, Mendelian randomization uses single nucleotide polymorphisms (SNPs), identified via GWAS, as instrumental variables to test for causality between traits. In doing so, Mendelian randomization takes advantage of the fact that SNP genotypes are randomly allocated during gamete formation (Mendel’s second law) and are thus generally not susceptible to the reverse causation and confounding that makes results from classical observational studies difficult to interpret [11-15].

We are aware of two previous studies that used Mendelian randomization to investigate the relationship between bipolar disorder and BMI. The first, by Hartwig et al., found no evidence that BMI is causal for bipolar disorder, and did not investigate whether bipolar disorder is causal for BMI [14]. The second, by Coleman et al., also found no evidence that BMI is causal for BD, but reported a nominally, yet not statistically, significant negative relationship in the direction from bipolar disorder to BMI [16]. The fact that Hartwig et al. did not investigate the impact of bipolar disorder on BMI and that Coleman et al. failed to find a statistically significant effect, may, however, reflect the relatively small sample sizes of the bipolar disorder GWAS providing data for these two studies. Specifically, the analysis by Hartwig et al. was based on GWAS data from Sklar et al. from 2011 [17] involving 7481 patients with bipolar disorder and 9250 controls, while the analysis by Coleman et al. was based on GWAS data by Stahl et al. from 2019 [18] involving 20,352 patients with bipolar disorder and 31,358 controls. More recently, Mullins et al. [19] have conducted a substantially larger GWAS, involving 41,917 patients with bipolar disorder and 371,549 controls. Therefore, with the aim of casting further light on the nature of the association between bipolar disorder and BMI/obesity, we conducted a bidirectional two-sample mendelian randomization study using summary statistics from the Mullins et al. [19] GWAS of bipolar disorder and from a UK Biobank GWAS of BMI [20].

## Methods

### Summary statistics for bipolar disorder

For the bipolar disorder instrumental variable, we used summary statistics from the GWAS by Mullins et al.[19], which was based on 41,917 patients with bipolar disorder and 371,549 ancestry-matched controls. The patients with bipolar disorder were diagnosed in accordance with DSM-IV, ICD-9 or ICD-10 using a standardized approach involving either trained interviewers, clinician-administered checklists or medical record review. Approximately 80% of the patients had type 1 bipolar disorder, with the remainder having type 2. The summary statistics (results from single-SNP regression) were retrieved from the homepage of the Psychiatric Genetics Consortium: https://www.med.unc.edu/pgc/download-results/.

### Summary statistics for BMI

For the BMI genetic instrumental variable, we use data derived from the UK Biobank [20]. The UK Biobank has genotyped approximately 500.000 individuals aged 40-69 years, who have also been phenotyped in a number of aspects, including calculation of BMI. For the primary analysis of the present study, we used summary statistics for BMI computed by the Neale Lab (available at: http://www.nealelab.is/uk-biobank). For secondary analyses, we used summary statistics for fat mass, fat-free mass and height from the same source. These summary statistics are based on data from 359,983 (BMI), 354,244 (fat mass), 354,808 (fat-free mass) and 360,388 (height) individuals, respectively.

### Data preparation and quality control

For bipolar disorder, we excluded SNPs with sample size <370,000, while for BMI and its components, we excluded SNPs that had an information score <0.95 (note that information scores were not available for bipolar disorder, while per-SNP sample sizes were not available for BMI and its components). We also excluded SNPs with alleles A & T or C & G (to avoid strand alignment errors) or that were not present in the 1000 Genome Project data [21]. Finally, to generate the genetic instrumental variables to be used in the Mendelian randomization analyses (bipolar disorder, BMI, fat mass, fat-free mass and height), we identified the SNPs that passed the threshold for genome-wide significance (P<5e-8) for these traits and subsequently carried out thinning for each trait until no SNP pairs remained within 3llJcM with correlation squared >0.05 (using SNP-SNP correlations computed across the European samples in the 1000 Genome Project data [21]. Figure 1 illustrates the distribution of the genetic instrumental variable scores for BD and BMI.

**Figure 1.**
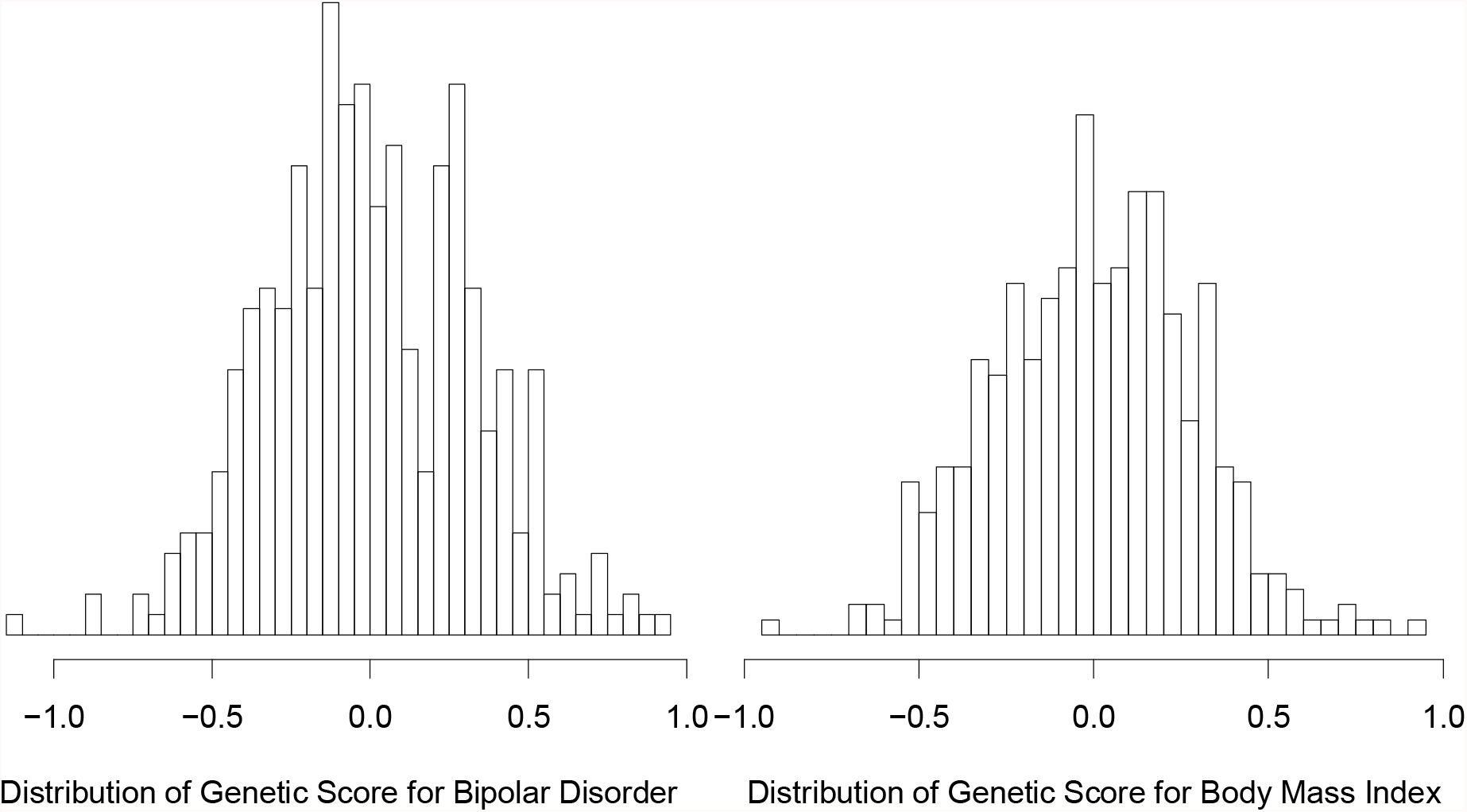
Illustration of the genetic scores for bipolar disorder and BMI. This figure shows the distribution of the genetic scores for BD (left) and BMI (right), across 404 non-Finnish European individuals from the 1000 Genome Project. The genetic scores for BD are computed using 53 SNPs, while those for BMI are computed using 517 SNPs.

### Bidirectional Mendelian randomization analyses

The analyses were carried out using the R package “MendelianRandomization” [22]. For each exposure-outcome pair, we used three types of MR analyses: inverse variance weighted (IVW) regression, weighted median regression and Egger regression. Each of these analyses measures the strength of causal relationship via the regression slope (for example, a slope of one indicates that increasing the exposure by one unit causes the outcome to increase by one unit, while a slope of zero indicates no causal relationship). IVW regression estimates the slope assuming no horizontal pleiotropy (i.e., that SNPs causal for the exposure are not also causal for the outcome). Weighted median regression instead assumes that at most 50% of the SNPs are pleiotropic (strictly, that at most 50% of the information comes from pleiotropic SNPs). Finally, Egger regression estimates the slope allowing for directional pleiotropy (i.e., that any pleiotropy effects are consistent across SNPs [12]). Our primary MR analysis performs two tests (i.e., whether bipolar disorder affects BMI and whether BMI affects bipolar disorder), and the Bonferroni-adjusted threshold for statistical significance was therefore 0.05/2=0.025. Subsequently, having determined that genetic liability for bipolar disorder seems to be causal for BMI, we explored which of the three components of BMI, namely fat mass, fat-free mass and height, is affected by the genetic liability to bipolar disorder using the same methods as described for the primary analyses, including the same genetic instrumental variable for bipolar disorder.

## Results

Table 1 describes the summary statistics for bipolar disorder and BMI. The genetic instrumental variable for bipolar disorder contained 53 SNPs and explained 0.5% of phenotypic variance, while the genetic instrument for BMI contained 517 SNPs and explained 7.1% of phenotypic variance.

**Table 1.** Details of summary statistics for bipolar disorder and BMI.

|  | **Bipolar disorder** | **BMI** |
| --- | --- | --- |
| Number of SNPs after quality control | 4371368 | 6183788* |
| Number of SNPs in common | 4347709** |  |
| Number of independent genome-wide significant ($P < 5e-8$) SNPs | 53 | 517 |
| Phenotypic variance explained by significant SNPs | .005 | .071 |

\* For the secondary analysis on fat mass, fat free mass and height, the corresponding numbers were 6183721, 6183765 and 6183812, respectively. \*\* For the secondary analysis on fat mass, fat free mass and height, the corresponding numbers were 4347716, 4347711 and 4347704, respectively.

The results from the primary MR analysis are reported in Table 2 and Figure 2. We found that genetic liability to bipolar disorder reduces BMI (the slope p-values from IVW regression, weighted median regression and Egger regression are 0.009, 1.3e-6 and 0.004, respectively). Given that the intercept from the Egger regression is different from zero (P=0.019), we believe the estimated slope from Egger regression is most reliable as it is computed allowing for directional pleiotropy. This estimate indicates that a one unit increase in the log odds ratio for developing bipolar disorder reduces BMI by approximately 0.2 standard deviations (SD). As the SD of BMI is 4.7m/kg^2^, this indicates that doubling the genetic liability to bipolar disorder (i.e., increasing the log odds by 0.69), on average reduces BMI by 0.6 kg/m2. When we reversed the direction of analysis, we found no evidence that BMI is causally linked to the risk of developing bipolar disorder (the slope p-values from IVW regression, weighted median regression and Egger regression are 0.592, 0.486 and 0.714, respectively).

**Table 2.**
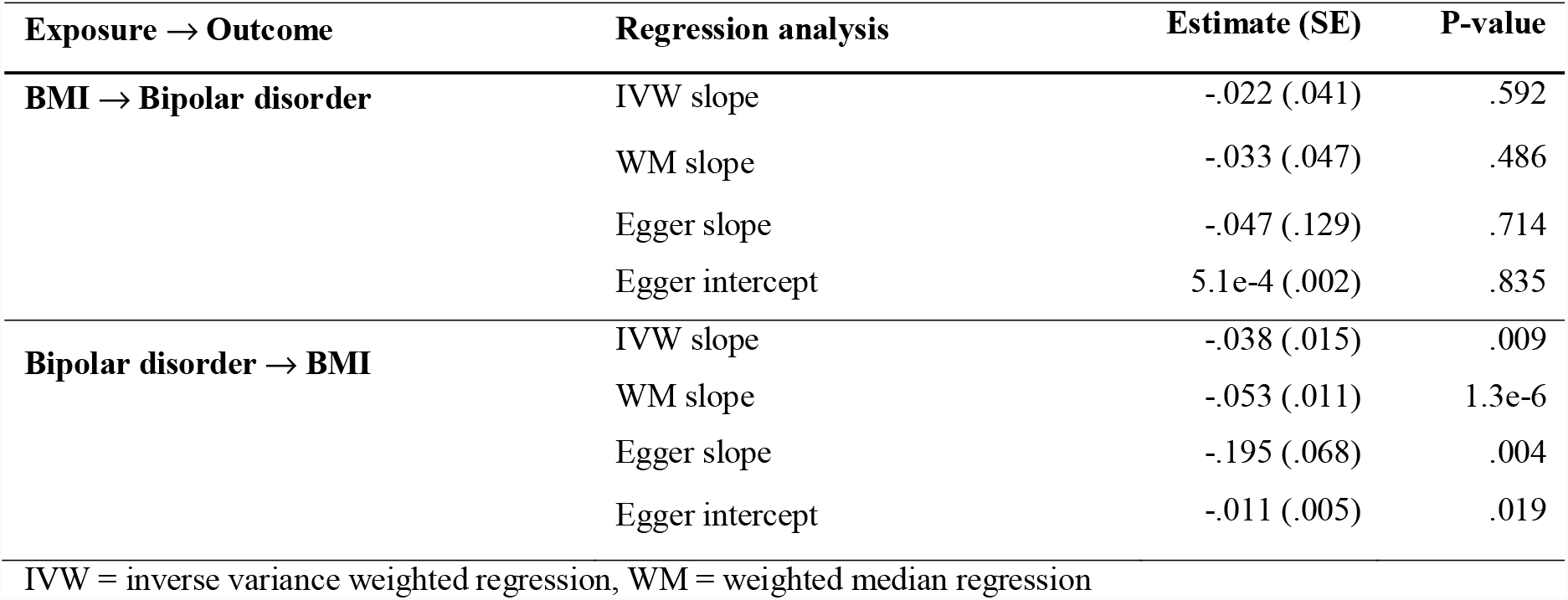
Tests of the causality of the relationship between bipolar disorder and BMI.

**Figure 2.**
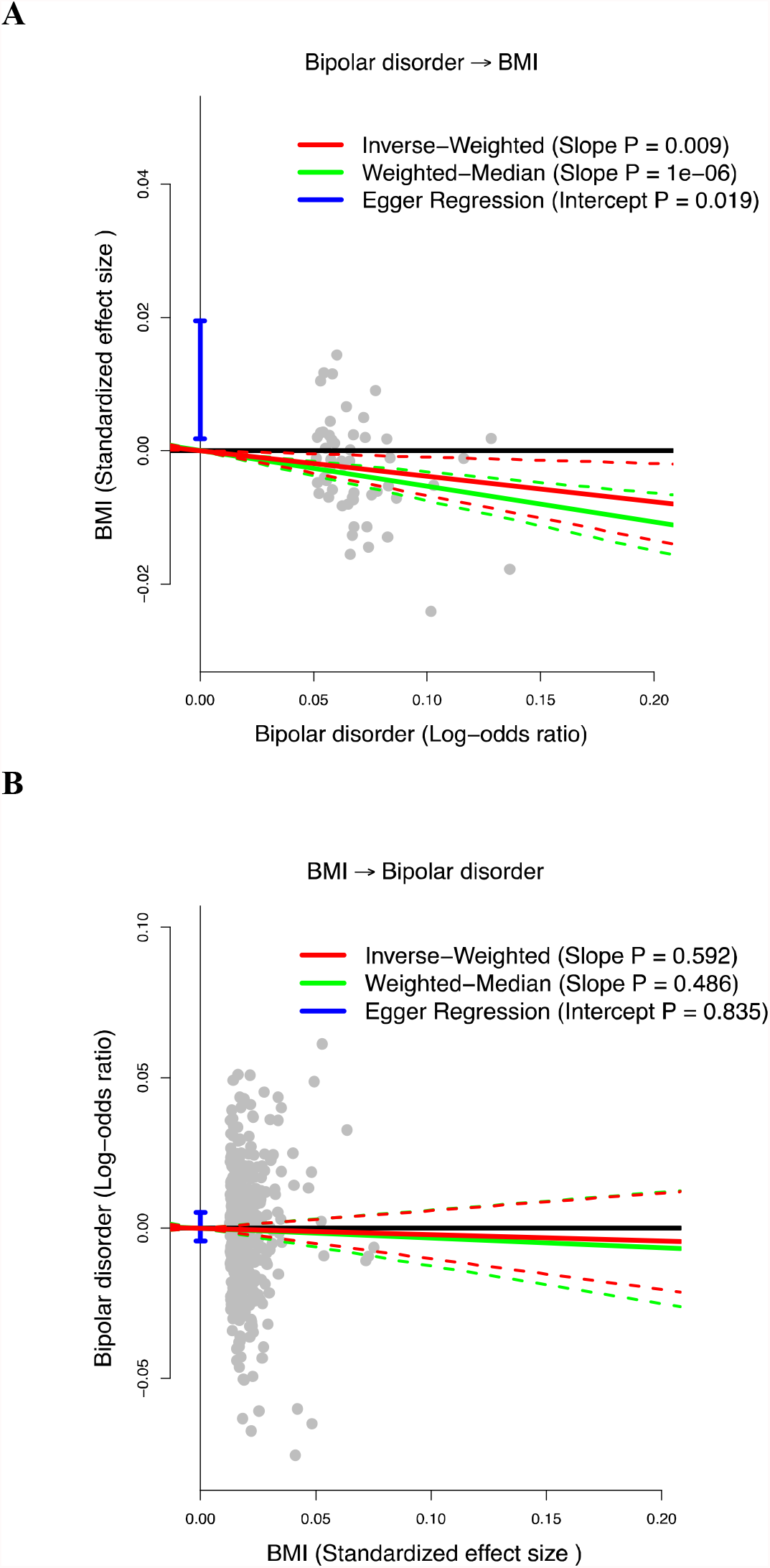
Tests of the causality of the relationship between bipolar disorder and body mass index. A. Displays results from the test of whether bipolar disorder is causal for BMI. B. Displays results from the test of whether BMI is causal for bipolar disorder. In each plot, grey points report effect sizes for the independent genome-wide significant SNPs for the exposure (for bipolar disorder, the units are log-odds ratios; for BMI, the units are standardized effects). The solid (dashed) red lines mark the slope (95%CI) from inverse variance weighted regression. The solid (dashed) green lines mark the slope (95%CI) from weighted median regression. The horizontal (vertical) blue lines mark the slope (95%CI for the intercept) from Egger regression.

The results of the secondary analysis focusing on bipolar disorder and the components of BMI, are shown in Table 3. We found that genetic liability to bipolar disorder reduces fat mass (the slope p-values from IVW regression, weighted median regression and Egger regression are 0.005, 1.0e-5 and 0.004 respectively), mixed evidence that genetic liability to bipolar disorder reduces fat-free mass (the slope p-values from IVW regression, weighted median regression and Egger regression are 0.037, 2.1e-6 and 0.061, respectively), and no evidence that genetic liability to bipolar disorder is causal for height (the slope p-values from IVW regression, weighted median regression and Egger regression are 0.410, 0.099 and 0.759, respectively).

**Table 3.**
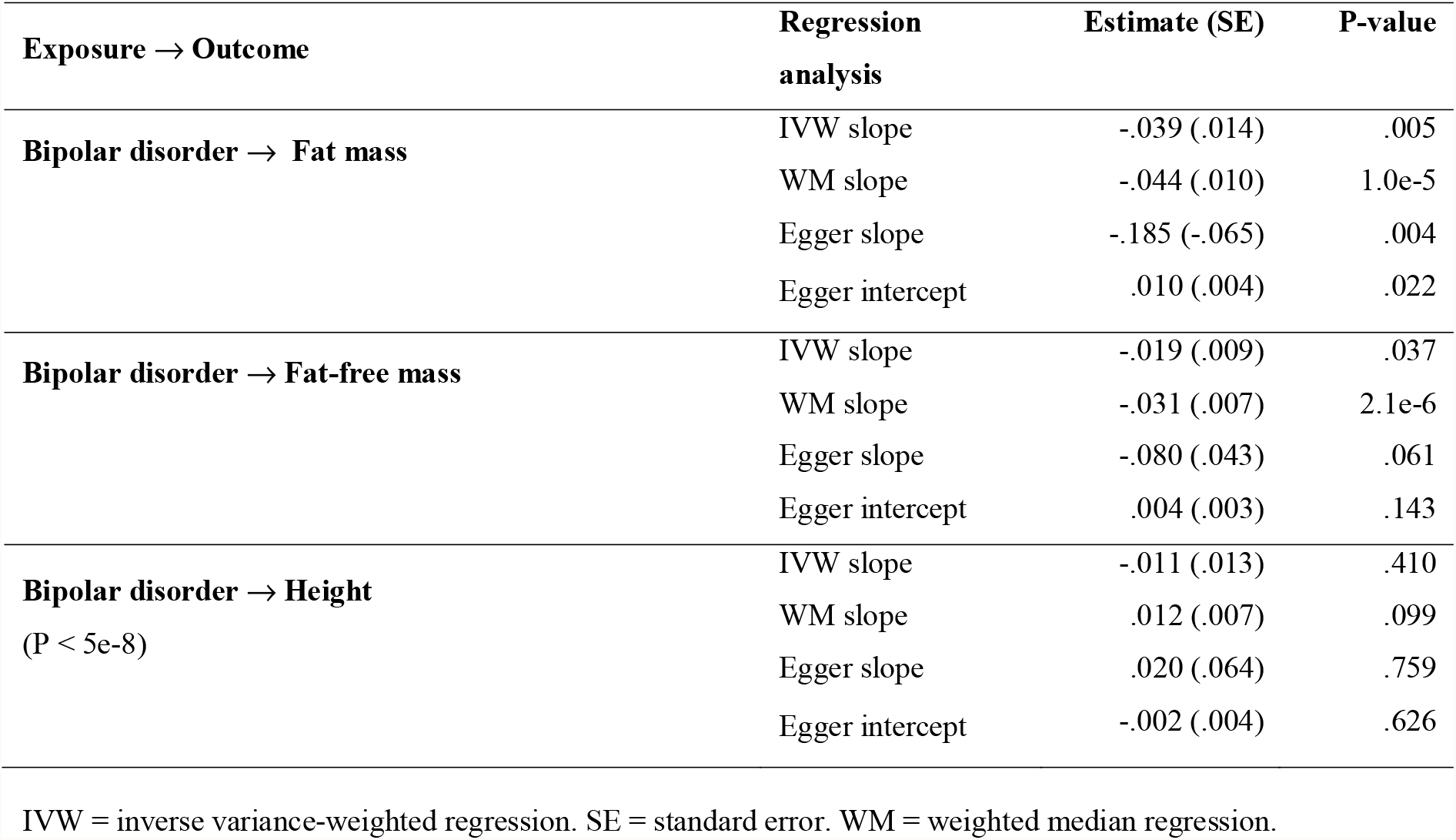
Tests of the causality of the relationship between bipolar disorder and fat mass, fat-free mass and height.

## Discussion

In this bidirectional Mendelian randomization study, we sought to determine the causal relationship between the genetic liability to bipolar disorder and BMI. The results show that the higher the genetic liability for bipolar disorder, the lower the BMI. The findings from the analyses stratified on the components of BMI (fat mass, fat-free mass and height) suggest that this is likely due to genetic liability for bipolar disorder leading to reduced fat mass. Conversely, we found no evidence that BMI is causally linked to the risk of developing bipolar disorder.

The results of this study corroborate the preliminary, but inconclusive, findings from the analysis carried out by Coleman et al. [16], which was based on data from a smaller GWAS than the one used in this study, and consequently had a less powerful genetic instrumental variable (13 SNPs instead of 53 SNPs) [15]. This reflects the fact that the power of a Mendelian randomization study will increase as the size of the GWAS grows, and the resulting instrumental variables explain increasing proportions of phenotypic variance.

Our findings may be hard to reconcile with experience from clinical practice as well as the many studies documenting high BMI/obesity rates among individuals with bipolar disorder [6]. Here it is, however, important to keep in mind that these studies have examined BMI/obesity in the context of *manifest* bipolar disorder (dichotomously defined), whereas the Mendelian randomization analysis uses the *genetic liability to* bipolar disorder (a continuous score – see Figure 1) as exposure. This distinction is crucial, because manifest bipolar disorder typically requires treatment with mood-stabilizing drugs, many of which have weight gain as a prominent side effect [23-26]. In the present study, the continuous instrumental variable based on the genetic liability to bipolar disorder is employed in the UK Biobank in which the proportion of individuals with bipolar disorder (requiring pharmacological treatment) is quite low [27], but yet probably covers a very large proportion of the continuous spectrum going from no genetic liability to bipolar disorder, to very high genetic liability to bipolar disorder. Taken together, this goes to suggest that the high BMI/obesity rate among individuals with bipolar disorder is not a direct consequence of the genetic liability to bipolar disorder, but rather attributable to factors that more exclusively affect those with manifest bipolar disorder, such as side effects of pharmacological treatment [23-26] poor diet [28, 29] and sedentary life style [30]. In support of this explanation, in their paper entitled “Weight gain occurs after onset of bipolar illness in overweight bipolar patients”, Shah et al. found that the mean remembered weight at the age of 18 years was numerically lower (albeit not statistically significantly so) among 32 patients with bipolar disorder (mean BMI of 30.3 at the time of the study) compared to 32 matched healthy controls (mean BMI of 24.3 at the time of the study) [18]. Along the same lines, based on data from a substantially larger sample of patients with bipolar disorder (n=3,115), Tohen et al. demonstrated that those with a first episode mania had lower BMI compared to patients with multiple episodes of mania (BMI of 24.6 vs. 26.3, p<0.001) [19].

While the arguments above provide a reason why many individuals with manifest bipolar disorder are overweight even though this is not directly genetically driven, it does, however, not explain why increased genetic liability to bipolar disorder leads to reduced BMI (and fat mass) as suggested by this study. With the symptoms of the opposing mood poles of bipolar disorder in mind [7-9], we speculate that both motor hyperactivity associated with subthreshold (hypo)mania, and/or decreased appetite in relation to subthreshold depression could be potential mediators of the observed association. Furthermore, it has been demonstrated that there is a positive correlation between the genetic risk for bipolar disorder and school grades/educational attainment [31, 32], which may imply that the genetic susceptibility to bipolar disorder also leads to increased health literacy and, in turn, to better diet and more exercise [33].

From a clinical perspective, the findings of this study are constructive in the sense that they suggest that the weight problems/obesity commonly observed in bipolar disorder may not be as “intrinsic” as could perhaps be expected. Conversely, they are likely driven by correlates of manifest bipolar disorder, such as side effects of pharmacological treatment, poor diet and sedentary lifestyle that are all (at least theoretically) modifiable and can be targeted as part of clinical management. Unfortunately, the evidence base for management of obesity in bipolar disorder is virtually absent [34].

There are some limitations to this study, which should be taken into account. First and foremost, Mendelian randomization relies upon three key assumptions: (i) the instrumental variables (i.e., the SNPs) are causal for the exposure variable, (ii) the instrumental variables are not associated with confounders of the exposure-outcome association, (iii) the instrumental variables only affect the outcome through the exposure variable (i.e. there is no horizontal pleiotropy). We are confident that (i) and (ii) hold true because we considered only SNPs robustly associated with the exposure (P<5e-8), and because SNPs are randomly allocated during meiosis. With regard to (iii), when testing whether the genetic liability to bipolar disorder was causal for BMI, we found evidence of pleiotropy, potentially invalidating our results from the IVW regression. However, we note that the causal relationship remained significant when instead using weighted median regression (which allows for up to half the instrumental variables to exhibit pleiotropy) and also when using Egger Regression (which allows for directional pleiotropy). Second, we note that some samples were common to both the bipolar disorder and the BMI GWAS (there were 1454 individuals with bipolar disorder and 58,113 controls from the UK Biobank in the GWAS of bipolar by Mullins et al.[19]). Although this overlap is likely to have had a very minor impact [35], the Mendelian randomization analysis should ideally be repeated using data from fully independent GWAS, and when results from larger GWAS (of bipolar disorder in particular), become available. Third, the measurements of fat and fat-free mass from the UK Biobank were obtained using bioelectrical impedance analysis on the Tanita BC418MA body composition analyzer, which is considered to be less accurate than e.g., dual-energy x-ray absorptiometry (DEXA). However, in the present study, the level of accuracy in distinguishing between fat and fat-free mass appears to have been sufficient to show that the genetic liability to bipolar disorder is predominantly affecting BMI via reduction of fat mass.

In conclusion, the findings of this study strongly suggest that the increased BMI observed among individuals with bipolar disorder does not stem directly from the genetic liability to bipolar disorder, but is rather a product of downstream consequences of manifest bipolar disorder, such as side effects to pharmacological treatment, poor diet and sedentary lifestyle, that are modifiable and may be targeted as part of clinical management.

## Data Availability

The GWAS summary statistics can be downloaded from the Psychiatrics Genomics Consortium (https://www.med.unc.edu/pgc/download-results) and from the Neale Lab (http://www.nealelab.is/uk-biobank). Genetic data from the 1000 Genomes Project can be downloaded from the project's website (https://www.internationalgenome.org).

## References

1. Laursen, T.M., Life expectancy among persons with schizophrenia or bipolar affective disorder. Scizophrenia Research, 2011. Volume 131, Issues 1–3.

2. Kessing, L.V., Vradi, E., & Andersen, P. K., Life expectancy in bipolar disorder. Bipolar Disord., 2015. 17(5): p. 543–548.

3. Xu, H., Cupples, L. A., Stokes, A., & Liu, C.-T., Association of Obesity With Mortality Over 24 Years of Weight History: Findings From the Framingham Heart Study. JAMA Netw Open, 2018. 1(7): p. e184587.

4. Westman, J., Hällgren, J., Wahlbeck, K., Erlinge, D., Alfredsson, L., & Ösby, U, Cardiovascular mortality in bipolar disorder: a population-based cohort study in Sweden. BMJ, 2013. 3(4): p. e002373.

5. Weiner M, Warren L., Fiedorowicz JG., Cardiovascular Morbidity and Mortality in Bipolar Disorder. Ann Clin Psychiatry, 2011. 23(1): p. 40–47.

6. Liu YK, Ling .S., Lui LMW, et al., Prevalence of type 2 diabetes mellitus, impaired fasting glucose, general obesity, and abdominal obesity in patients with bipolar disorder: A systematic review and meta-analysis [published online ahead of print, 2021 Dec 26]. J Affect Disord., 2021. 300:449–461.

7. World Health Organization, The ICD-10 classification of mental and behavioural disorders. Diagnostic criteria for research. Geneva: WHO. 1993.

8. American Psychiatric Association, Diagnostic and Statistical Manual of Mental Disorders, 4th Edition, Text Revision. Washington, DC. 2000..

9. American Psychiatric Association, Diagnostic and Statistical Manual of Mental Disorders, 5th Edition. Washington, DC. 2013..

10. Cross-Disorder Group of the Psychiatric Genomics Consortium, L.S., Ripke S, et al, Genetic relationship between five psychiatric disorders estimated from genome-wide SNPs. nat genet, 2013. 45(9): p. 984–994.

11. Speed MS J.O., Børglum AD, Speed D, Østergaard SD, Investigating the Association Between Body Fat and Depression via Mendelian Randomization Transl Psychiatry, 2019. 9: p. 184.

12. Bowden J, Davey Smith G., Burgess S, Mendelian randomization with invalid instruments: effect estimation and bias detection through Egger regression. International Journal of Epidemiology, 2015. 44(2): p. 512–525.

13. Bowden J, Davey Smith G., Haycock PC, Burgess S., Consistent Estimation in Mendelian Randomization with Some Invalid Instruments Using a Weighted Median Estimator. Genet Epidemiol, 2016. 40(4): p. 304–314.

14. Davey Smith G, Ebrahim S., ‘Mendelian randomization’: can genetic epidemiology contribute to understanding environmental determinants of disease?. International Journal of epidemiology, 2003. 32(1): p. 1–22.

15. Speed D, Hemani G., Speed MS; Major Depressive Disorder Working Group of the Psychiatric Genomics Consortium, Børglum AD, Østergaard SD, Investigating the causal relationship between neuroticism and depression via Mendelian randomization. Acta Psych Scand., 2019. 139(4): p. 395–397.

16. Coleman JRI, Gaspar HA., Bryois J; Bipolar Disorder Working Group of the Psychiatric Genomics Consortium; Major Depressive Disorder Working Group of the Psychiatric Genomics Consortium, Breen G. The Genetics of the Mood Disorder Spectrum: Genome-wide Association Analyses of More Than 185,000 Cases and 439,000 Controls. Biol Psychiatry., 2020. 88(2):169-184.

17. Psychiatric GWAS Consortium Bipolar Disorder Working Group. Fullerton, Janice M [added]; Hyoun, Phil L [corrected to Lee, Phil H]; Meng, Fan Guo [corrected to Meng, Fan]], Large-scale genome-wide association analysis of bipolar disorder identifies a new susceptibility locus near ODZ4 Nat Genet., 2011. 43(10):977-983..

18. Stahl EA, Breen G., Forstner AJ, McQuillin A, Ripke S, Trubetskoy V, et al., Genome-wide association study identifies 30 loci associated with bipolar disorder. Nat Genet., 2019 51(5).

19. Mullins N, Forstner AJ., O’Connell KS, et al., Genome-wide association study of more than 40,000 bipolar disorder cases provides new insights into the underlying biology. Nature Genetics, 2021. 53(6): p. 817–829.

20. Bycroft C, Freeman C., Petkova D, et al., The UK Biobank resource with deep phenotyping and genomic data. Nature, 2018. 562(7726): p. 203–209.

21. 1000 Genomes Project Consortium, Auton A., Brooks LD, et al., A global reference for human genetic variation. Nature., 2015. 526(7571): p. 68–74..

22. Yavorska OO, Burgess S., MendelianRandomization: an R package for performing Mendelian randomization analyses using summarized data. International Journal of Epidemiology, 2017. 46(6): p. 1734–1739.

23. Köhler-Forsberg O, Gasse C., Hieronymus F, et al., Pre-diagnostic and post-diagnostic psychopharmacological treatment of 16 288 patients with bipolar disorder. Bipolar Disord., 2021. 23(4): p. 357–367..

24. Bjørklund L, Hordal HT., Mors O, Østergaard SD, Gasse C., Trends in the psychopharmacological treatment of bipolar disorder: a nationwide register-based study. Acta Neuropsychiatr., 2016. 28(2)(75–84.).

25. Patino LR, Klein CC., Strawn JR, et al., A Randomized, Double-Blind, Controlled Trial of Lithium Versus Quetiapine for the Treatment of Acute Mania in Youth with Early Course Bipolar Disorder. J Child Adolesc Psychopharmacol., 2021. 31(7)(485–493.).

26. Solmi M F.M., Ostinelli EG, et al., Safety of 80 antidepressants, antipsychotics, anti-attention-deficit/hyperactivity medications and mood stabilizers in children and adolescents with psychiatric disorders: a large scale systematic meta-review of 78 adverse effects.. World Psychiatry., 2020. 19(2)(214–232).

27. Davis KAS, Ccoleman JRI., Adams M, et al., Mental health in UK Biobank - development, implementation and results from an online questionnaire completed by 157 366 participants: a reanalysis. 2020, BJPsych Open.. 6(2)(e18.).

28. Jacka FN, Pasco JA., Mykletun A, et al., Diet quality in bipolar disorder in a population-based sample of women. J Affect Disord., 2011. 129(1-3):: p. 332–337..

29. Teasdale SB, Ward PB., Samaras K, et al., Dietary intake of people with severe mental illness: systematic review and meta-analysis. Br J Psychiatry., 2019. 214(5)(251–259.).

30. Vancampfort D, Firth J., Schuch FB, et al., Sedentary behavior and physical activity levels in people with schizophrenia, bipolar disorder and major depressive disorder: a global systematic review and meta-analysis. World Psychiatry., 2017. 16(3): p. 308–315.

31. Østergaard, S.D., McGrath, J. J., Mors, O., Mortensen, P. B., & Petersen, L. V., Polygenic risk score for bipolar disorder and school grades. Journal of Affective Disorders, (2020) 263.

32. Brainstorm Consortium, Antilla V., Bulik-Sullivan B, et al., Analysis of shared heritability in common disorders of the brain. Science., 2018. 360(6395)(eaap8757.).

33. Friis K, Lasgaard M., Rowlands G, Osborne RH, Maindal HT., Health Literacy Mediates the Relationship Between Educational Attainment and Health Behavior: A Danish Population-Based Study. J Health Commun., 2016. 21(Sup2):: p. 54–60.

34. Tully A, Smyth S., Conway Y, et al., Interventions for the management of obesity in people with bipolar disorder. Cochrane Database Syst Rev., 2020. 7(7)(CD013006).

35. Burgess S, Davies NM., Thompson SG., Bias due to participant overlap in two-sample Mendelian randomization. Genet Epidemiol., 2016. 40(7): p 597–608.

